# The Effect of Perceived Social support on Cervical Cancer Screening among Nepali women residing in Dhulikhel and Banepa municipality

**DOI:** 10.1101/2024.05.21.24307693

**Authors:** Gaël Cruanes, Pratiksha Paudel, Urja Regmi, Darlene R House, Archana Shrestha, Bandana Paneru

## Abstract

**Background:** Cervical cancer ranks as the most common cancer among Nepalese women with a high incidence and mortality. Despite evidence that effective screening programs reduce disease burden, screening services are under-utilized. In various other countries, perceived social support has been shown to increase cervical cancer screening uptake among women.

**Objectives:** This study assessed the association between perceived social support and cervical cancer screening uptake among women residing in semi-urban areas of Kavrepalanchok district (Dhulikhel and Banepa), Nepal.

**Methods:** We conducted a cross-sectional study among women aged 30-60 years using telephone interview method from 15^th^ June to 15^th^ October 2021. A validated Multidimensional Scale of Perceived Social Support (MSPSS) was utilized to measure perceived social support. We obtained information on cervical cancer screening uptake through self-reported responses. A Mann Whitney analysis was performed to assess the association between perceived social support and cervical cancer screening uptake.

**Results:** Four hundred twenty-six women participated in the study. There was a significant association between significant others’ social support (p-value=0.007, 95% CI: 0.08-0.53) and family social support (p-value=0.04, 95% CI: 0.003-0.4).

**Conclusion:** Women residing in semi-urban areas of Nepal who had higher perceived social support from their significant other and family were more likely to have been screened for cervical cancer. Community interventions fostering conversations about reproductive health between husbands and their wives may increase perceived social support and contribute to higher uptake of cervical cancer screening.

## Introduction

Cervical cancer is the fourth most common cancer for women worldwide and leads to around 342,000 deaths per year (2020). ^[1]^ 85% of the global burden of cervical cancer can be attributed to low and middle income countries (LMICs). ^[2]^ According to *Sankaranarayanan R et al.*, about a third of the global cervical cancer burden comes from Nepal, Sri Lanka, and India. ^[3]^ In Nepal, cervical cancer is the most common cancer, with about 1,493 deaths occurring annually in 2020. ^[4]^ There are numerous barriers towards receiving cervical cancer screening in Nepal such as the cost of the screening, cancer stigma, fear of positive results, lack of test awareness, lack of knowledge around the disease, reluctance to screen, low-risk perception, and lack of time from the women. ^[5][6]^ It is important to note that Nepal has a patriarchal social structure, in which only 57.4% of women are literate, compared with 75.1% of men. ^[7]^ Family acceptance and support was perceived as essential for them to act and to accept cervical cancer screening services. ^[8]^

Social support is the perception or reality that one is cared for by a social network. In the case of community health, there is growing evidence that having multiple links of social networks leads to increases in health screening. ^[9]^ According to a scoping review by *Williams-Brennan, Leslie, et al* looking at perceived social support in developing countries, researchers reported that women who participated in cervical cancer screening were encouraged to do so by their family, friends or spouse ^[10, 11, 12, 13, 14, 15]^ and that married women were more likely to report having a Pap smear test performed than single women. ^[16, 17, 18, 19, 20, 21]^, ^[22]^ Additionally, in a study conducted in Sub-Saharan Africa, authors found that positive social interaction was significantly associated with Pap screening. ^[23]^ Comparatively in a study conducted in the United States, women with education less than or equal to high school diploma and those with high social support were 2.85 times (CI: 1.81, 4.49) more likely to be compliant with cervical cancer screening when compared to those with low social support. However, high social support did not make a difference for more educated women. ^[24]^ Similarly, talking to friends about issues related to health is associated with a more adequate frequency of having cervical cancer screening. ^[25]^ It is important to note that the studies in the scoping review were done in various populations around the world, including rural Mexico, rural Colombia, rural Argentina, northern Peru, Bulgaria, Romania, Taiwan, mainland China, Ghana and Northern India. There are no studies done in Nepal, where different social structures and cultural values may impact social support on cervical cancer screening.

Therefore, the objective of this study was to evaluate the association between perceived social support and cervical cancer screening uptake among women of semi-urban areas in central Nepal.

## Methods

### Study Design and Population

A community-based analytical cross-sectional survey was carried out among 429 women residing in Dhulikhel and Banepa municipality of Nepal who were eligible for cervical cancer screening. Dhulikhel and Banepa are ancient cities of Kavrepalanchok district, located about 30 kilometers east of the capital city (Kathmandu) with a total population of 39,047. ^[34]^

The eligibility criteria followed national cervical cancer screening guidelines, which meant they were not pregnant, had no previous history of cervical intraepithelial neoplasia (CIN) or cancer, and were between 30 to 60 years-old. The project has received IRB approval from the Ethical Review Board, Nepal Health Research Council (Jan 2020 /ref no 1699) and IRB at Icahn School of Medicine at Mount Sinai (Study-23-00362).

### Measures

#### Sociodemographics variables

The following sociodemographic variables were collected: age (in years), ethnicity (Brahmin, Chettri/Thakuri/Sanyasi, Newar, Magar/Rai/Limbu/Bhote/Sherpa/Tamang, Dalit), education (no formal education, primary education, secondary education, secondary and above), religion (Hindu, Non-Hindu), occupation (farmer, business, unemployed, others) and marital status (married and unmarried).

#### Perceived Social Support

To investigate perceived social support, the Multidimensional Scale of Perceived Social Support (MSPSS) was utilized. The MSPSS is a 12-item measure of perceived adequacy of social support from three sources: family, friends, & significant other; using a 5-point Likert scale (0 = strongly disagree, 5 = strongly agree). ^[35]^ When looking specifically at the role of the family in perceived social support, the scores from the 7 point likert scale on questions 3,4, 8 and 11 were summed and divided by 4. When looking specifically at the role of friends in perceived social support, the scores from the 7 point likert scale questions 6, 7, 9, and 12 were summed and divided by 4. The total score is a measure of overall perceived social support. A total score of 1 to 2.9 on MSPSS is considered to be low perceived social support. A total score of 3 to 5 on MSPSS is considered to be moderate perceived social support. A total score of 5.1 to 7 on MSPSS is considered to be high perceived social support. ^[35]^

### Cervical cancer screening

Cervical cancer screening uptake was assessed from the self-reported responses from an phone based interview to the question – “Has a healthcare worker ever tested you for cervical cancer?”

### Study protocol

This study was conducted in Dhulikhel and Banepa municipality of Kavrepalanchowk district, Nepal. Dhulikhel and Banepa are situated approximately 30 kilometers east of the capital city, Kathmandu. Banepa has a population of 55,628, while Dhulikhel Municipality has a population of 32,162, with an equal distribution of around 50% females in both areas. 17 government health facilities, 7 in Banepa and 10 in Dhulikhel municipality offer free-of-cost Visual Inspection with Acetic Acid (VIA). ^[36]^ Liquid-based cytology (LBC) and HPV testing are available at community-based hospitals.

### Selection and recruitment of participants

For the survey, we recruited 429 women, aged 30-60 years residing in Dhulikhel or Banepa municipality. Female Community Health Volunteers (FCHVs) played a critical role in identifying eligible women through home visits and informing them about our study. FCHVs serve as part of the Nepali community health workforce primarily supporting health education, counseling, outreach, and resource distribution. ^[37]^ Women aged 30-60 years were selected based on the cervical cancer screening target group as per the national guidelines for Cervical Cancer Screening and Prevention. ^[38]^ The inclusion criteria of the study were the following: (1) Women 30 to 60 years; (2) Uterus intact (3) Resident of Dhulikhel and Banepa. The exclusion criteria were (1) Pregnant; (2) Previous history of cervical intraepithelial neoplasia (CIN) or cancer. Research Assistants (RAs) recruited eligible and interested individuals to participate in the study and written informed consent was obtained from all participants.

### Survey data collection

At enrollment, the trained RAs had telephonic interviews with the participants using a standardized electronic questionnaire on kobotoolbox. The questionnaire assessed socioeconomic characteristics, including age, ethnicity, religion, occupation, marital status, education, cervical cancer screening and MSPSS. The interview lasted for around 50 to 60 minutes. For cervical cancer screening, the participants were instructed to use an HPV self-swab by a trained physician.

### Data Analysis

Categorical data were reported in frequency and percentage; and numerical data with means and standard deviation. Perceived social support was calculated using a five point Likert scale as mentioned earlier. We reported the difference in mean score among screened and unscreened groups using an unpaired t test and chi-squared test. All analyses were conducted using STATA version 13.0 (Stata Corp., College Station, Texas, USA) for cleaning, coding and statistical analysis.

## Results

Four hundred twenty nine women were surveyed and the average age of the participants surveyed was 41 years (SD 8 years). The majority of the study population was Brahmin/Chettri/Thakuri/Sanyasi (40.79%) and Newar (39.16%). In terms of education, The majority of the research population had secondary education (38.9%), followed by having no formal education) (29.6%). In terms of occupation, the majority of the population were farmers (40.8%) or unemployed (25.2%). The majority of participants were married 93%. The sociodemographic characteristics of the population are present in Table 1 below.

**Table 1:**
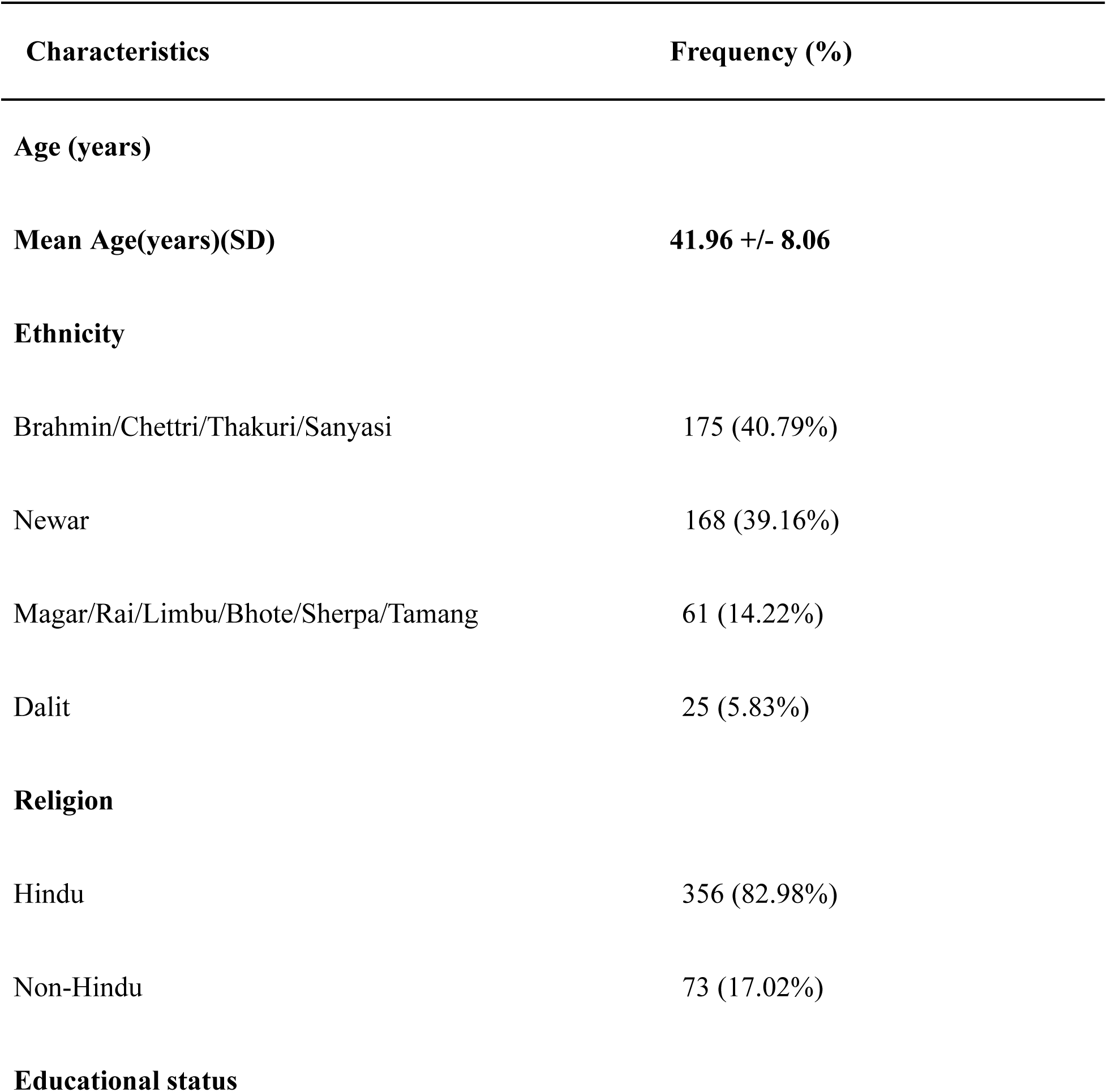
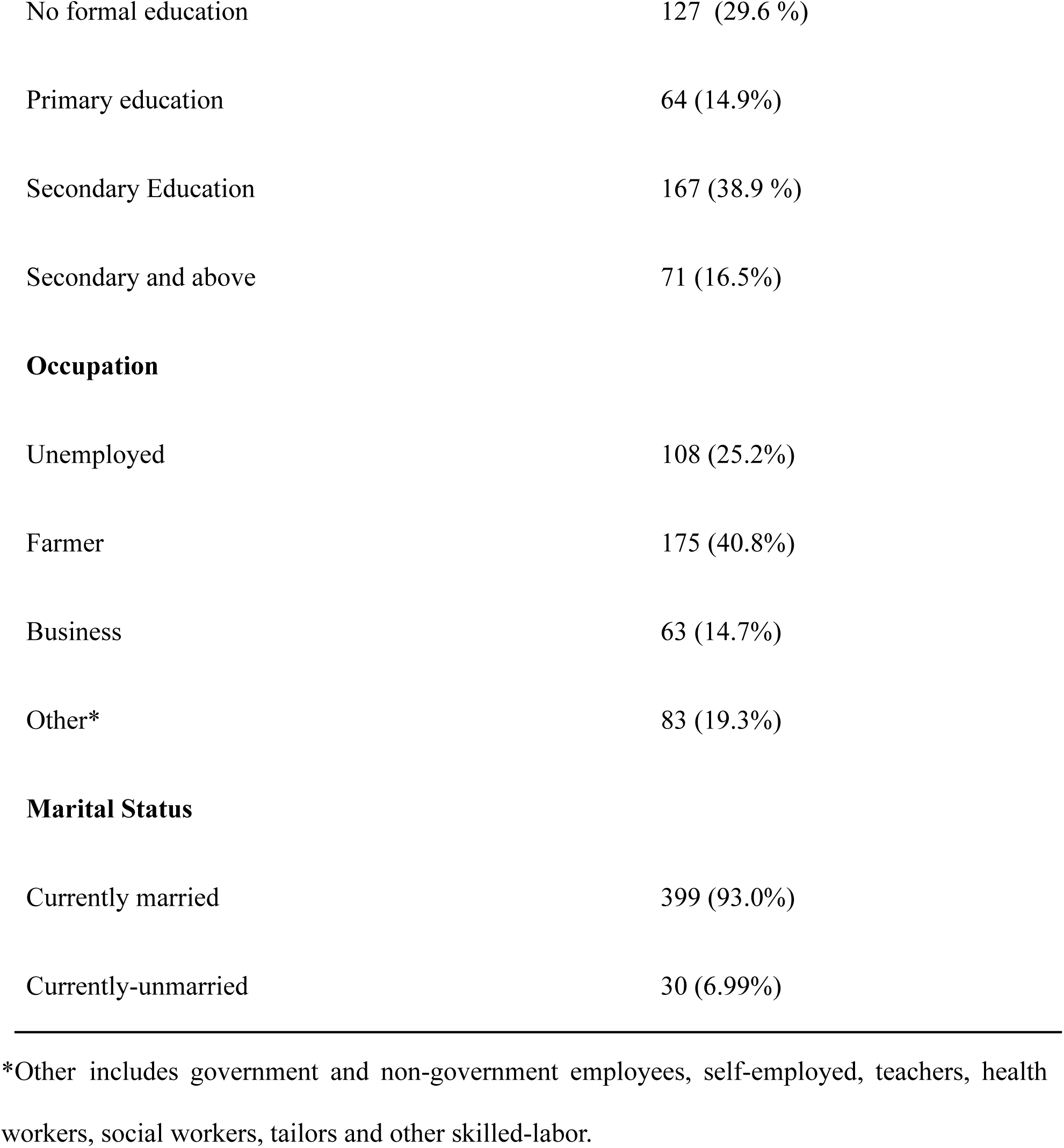
Sociodemographic Characteristics of Participants (n=429)

| **Characteristics** | **Frequency (%)** |
| --- | --- |
| **Age (years)** |  |
| **Mean Age(years)(SD)** | **41.96 +/- 8.06** |
| **Ethnicity** |  |
| Brahmin/Chettri/Thakuri/Sanyasi | 175 (40.79%) |
| Newar | 168 (39.16%) |
| Magar/Rai/Limbu/Bhote/Sherpa/Tamang | 61 (14.22%) |
| Dalit | 25 (5.83%) |
| **Religion** |  |
| Hindu | 356 (82.98%) |
| Non-Hindu | 73 (17.02%) |
| **Educational status** |  |
| No formal education | 127 (29.6 %) |
| Primary education | 64 (14.9%) |
| Secondary Education | 167 (38.9 %) |
| Secondary and above | 71 (16.5%) |
| **Occupation** |  |
| Unemployed | 108 (25.2%) |
| Farmer | 175 (40.8%) |
| Business | 63 (14.7%) |
| Other* | 83 (19.3%) |
| **Marital Status** |  |
| Currently married | 399 (93.0%) |
| Currently-unmarried | 30 (6.99%) |

\*Other includes government and non-government employees, self-employed, teachers, health workers, social workers, tailors and other skilled-labor.

Of the 429 women surveyed, 311 participants (72.5%) had never been screened for cervical cancer and 118 participants (27.51%) had been screened for cervical cancer.

According to Table 2 below, participants had high perceived overall social support. Perceived social support was also high according to subgroups of significant others, family and friends.

**Table 2:**
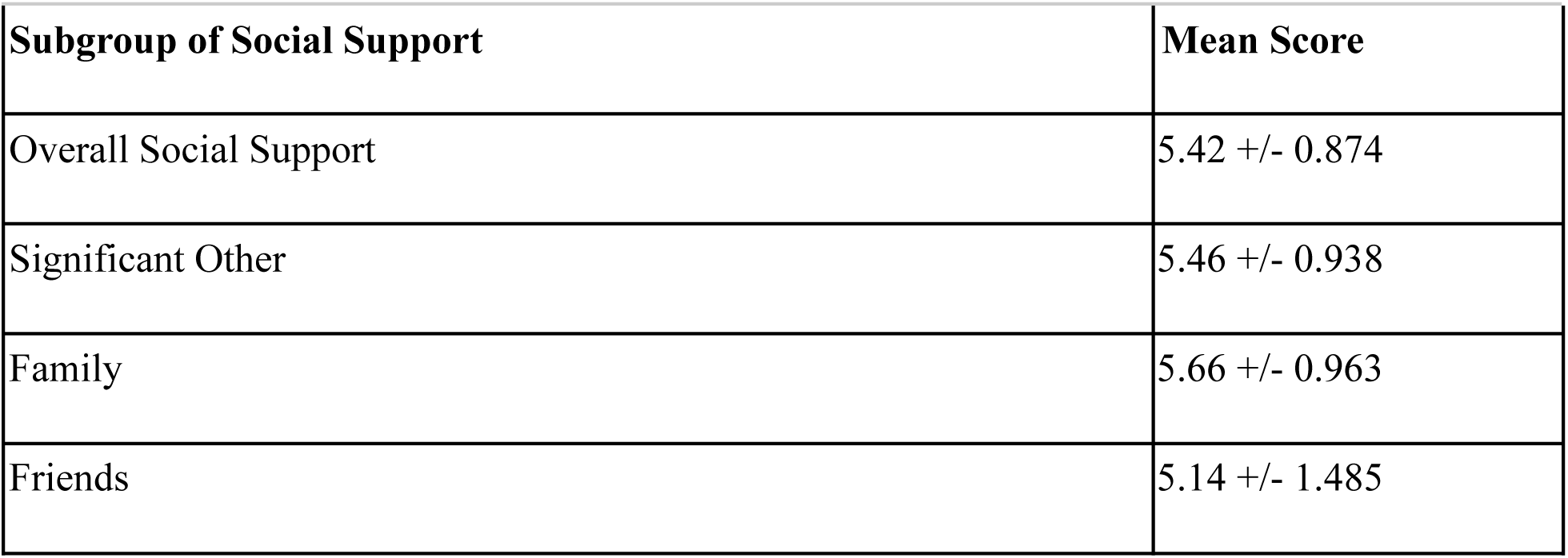
Mean Score of MSPSS Scale.

**Table 2:**
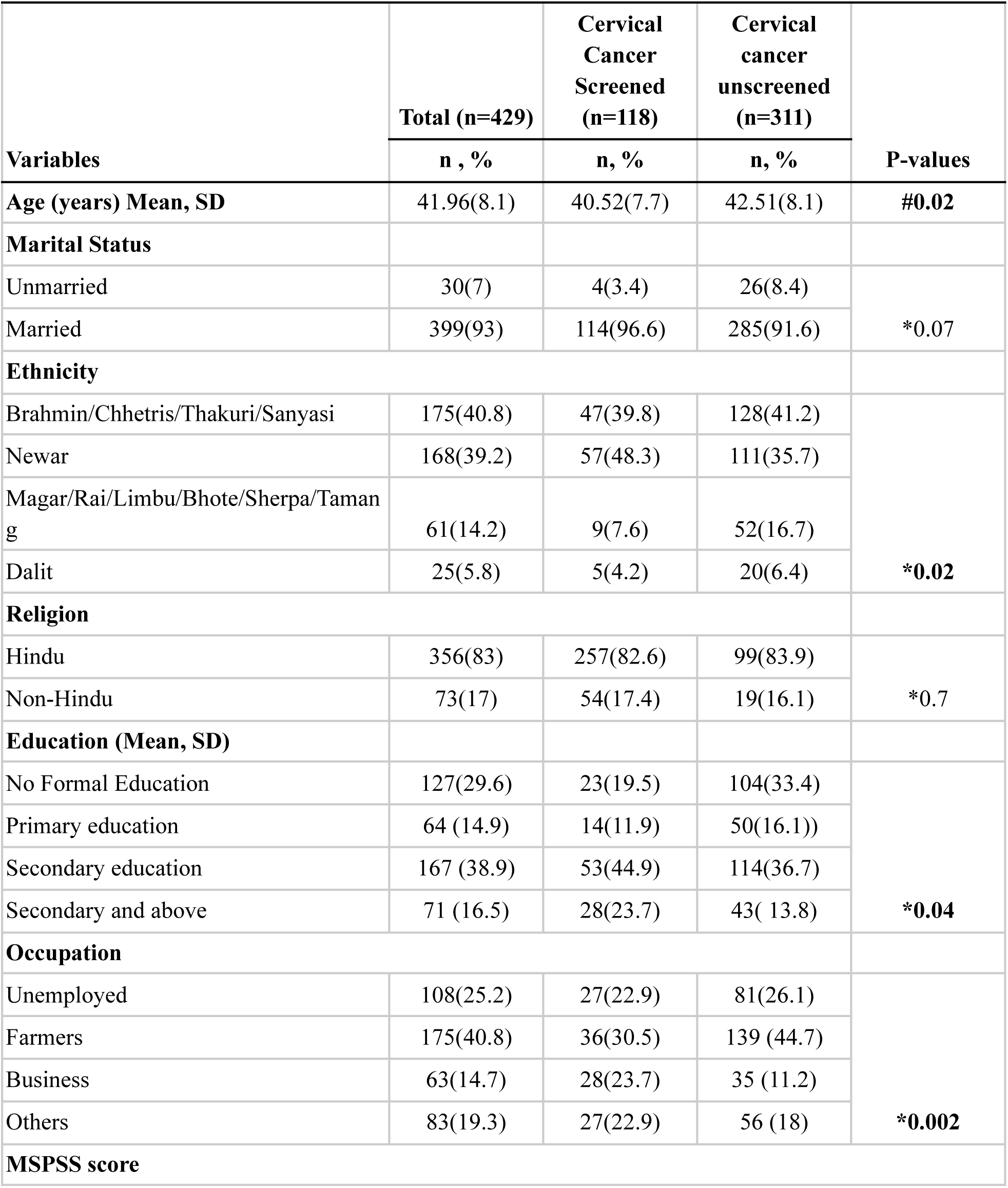
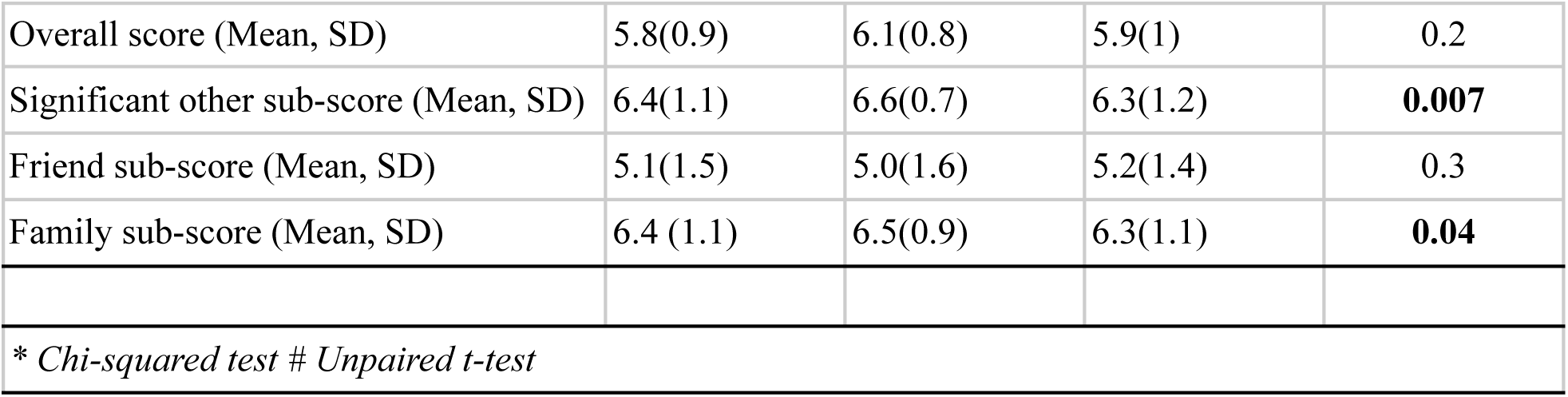
Factors associated with Cervical Cancer Screening.

We ran an unpaired t-test and chi square analysis to assess the association between the socio-demographic variables, social support and cervical cancer screening. There was a significant difference in mean age among screened and unscreened (p-value=0.02). Unscreened participants on an average were 1.98 years older as compared to screened. (95% CI: 0.28-3.69) Similarly, ethnicity was also significantly associated with screening (p-value=0.02). Participants that are of Dalit ethnicity had the lowest percentage of screening (4.2%), while Newar ethnicity had the highest screening percentage (48.3%). While comparing education and screening, secondary education had the highest percentage of screening 44.9% (p-value = 0.04) and 30% of those screened were farmers. (p-value=0.002)

While assessing the relation between perceived social support and cervical cancer screening, there was significant association between social support from significant others (p-value=0.007, 95%CI: 0.08-0.53) and social support from family (p-value=0.04, 95% CI: 0.003-0.4).

## Discussion

Perceived social support has a significant impact on cervical cancer screening uptake in semi-urban Nepal. Our study illustrates that when comparing subgroups of social support in cervical cancer uptake, the average score of significant other social support and family social support were found to be significantly higher among screened groups when compared to unscreened groups.

Our results are similar to results from a systematic review written by Akoto, Edem J., and Matthew J. Allsop. ^[39]^ Diminished support from a spouse can reduce the likelihood of engagement with screening. ^[40][41]^ The influence of a spouse extends to financial and emotional support, often sought from preparation through to treatment and follow-up stages. ^[42]^ Furthermore, where a spouse has a higher level of knowledge concerning the benefits of cancer screening, their spouse is more likely to participate in screening programs. ^[43]^ Among those who were screened, the most common reason for screening was health personnel’s advice (69.6%), whereas the least common reason was family’s advice (2.9%). ^[9]^ Our study gives credence towards the fact that family support is found to be more instrumental in women getting screened for cervical cancer.

Our results support findings from previous studies that show an association between social support and cervical cancer screening participation ^[10, 11, 12, 13, 14, 15 16, 17, 18, 19, 20, 21, 44, 45]^ These studies were done in a variety of low middle income countries around the world, including rural Mexico, rural Colombia, rural Argentina, northern Peru, Bulgaria, Romania, Taiwan, mainland China, Ghana and Northern India.

In May 2019, The WHO released a cervical cancer elimination initiative that set cervical cancer screening benchmarks for each country. WHO guidance set the guidance that 70% of women should be screened using a high-performance test by the age of 35, and again by the age of 45. ^[46]^ Our study reported that 27.51% had been screened for cervical cancer which unfortunately does not meet the WHO Standard that has been set for 2030. Comparatively, in India, the prevalence of cervical cancer screening was reported as 29.8% among women aged 30–49 years and 22.3% among women aged 15–49 years in 2015. ^[47][48]^ Comparatively, in Bangladesh, only 7.5% women aged between 30-49 group had gone under cervical screening in their lifetime according to Health Bulletin 2019. ^[49]^ Comparatively, in Bhutan, 90.8% of the eligible population was screened for cervical cancer screening of which 10,749 (9.2%) were HPV DNA positive. ^[50]^ This difference could be due to a multitude of factors including but not limited to cancer stigma, cultural stigma, fear of positive results, lack of test awareness, lack of knowledge around the disease, reluctance to screen, low-risk perception, and lack of time from the women. ^[5][6]^ Further work needs to be done to investigate the effectiveness of perceived social support Future studies and interventions should consider these social interactions and impact of social support on cervical cancer prevention program.

### Limitations

This study has a few limitations. Since this study is a cross-sectional study, temporality cannot be analyzed. The study was also conducted in two specific semi-urban settings so generalizability is limited to the rest of Nepal. Additionally, the survey responses were self reported so there is a chance of reporting bias or social desirability bias. Lastly, a multivariable analysis can not be run with the current parameters of the data.

## Conclusion

Nepali women who had perceived social support from significant others and family settings were more likely to have been screened for cervical cancer. Future public health awareness campaigns and efforts should be cognizant of this effect. These screening camps need to be mindful in targeting health behaviors while also not ignoring the influence of larger political, social, cultural and economic structures that contribute to receiving cervical cancer screening.

## Data Availability

All data produced in the present study are available upon reasonable request to the authors

## Acknowledgement

The authors would like to acknowledge and thank all the participants of the study as well as the female community health volunteers that helped us with outreach. We would also like to thank Dhulikhel Hospital and Dhulikhel Municipality for organizing visits with the community outreach centers, and aid in providing a platform for data collection.

## Conflict of Interests

The authors declare no conflict of interests

## Appendix

## Cervical Cancer Questions

Participants ID number________

Name of the interviewer________

**Eligibility Questions**

1. Do you have an intact uterus? Yes No
2. Do you have a previous history of cervical cancer? Yes No
3. Are you currently pregnant? Yes No
4. How old are you? Has to be between 30 to 60 years
5. Date of birth _______

**Consent Agree Disagree**

**Sociodemographic (Source: Dhulikhel Heart study)**

1. Full Name
2. Mobile Number
3. Ethnic Group

- Brahmin
- Chettri/Thakuri/Sanyasi
- Newar
- Magar/Tamang/Rai/Limbu
- Sherpa/Bhote
- Kami/Damai/Sarki/Gaaine/Baadi
- Others If others specify
4. Marital Status

- Married
- Unmarried
- Separated
- widowed
- Cohabiting
- Refused
5. Religion

- Hindu
- Buddhist
- Muslim
- Kirat
- Christian
- Others---specify
6. What is the highest grade or year of school you have ever completed, including college?
7. Which of the following describes your main status over the past 12 months?

- Government Employ
- Non Government Employ
- Self Employed
- Non Paid
- Homemaker
- Unemployed
- Student
- Farmer
- Teacher
- Health worker
- Business
- Others If others specify
8. Talking about the past year, what was your average earning?
9. Talking about the past year, what was your family’s annual earnings?
10. How many members are there in your family (share kitchen)? 11.

**Husband Information (sociodemographic including migration) (Source: Demographic Health Survey)**

1. How old was your husband on his last birthday?
2. What is the highest grade your husband completed?
3. What is your husband’s occupation? That is. What kind of work does he mainly do?

- Government Employ
- Non Government Employ
- Self Employed
- Non Paid
- Homemaker
- Unemployed
- Student
- Farmer
- Teacher
- Health worker
- Business
- Others If others specify?
4. Does your husband work away from home/ abroad? Yes No
5. How often does he come home?

- Once a week
- Once a month
- Once every 3 months
- Once in 6 months
- Once a year
- Once every 2 year
- More than 2 year
- Never come
6. When was the last time he came home?
7. How long did he stay?

- Days
- Weeks
- Months
- Years
- Others If others specify?

**Cervical cancer and screening**

1. Have you ever heard about Cervical cancer? Yes No don’t know
2. Where or from whom did you hear about cervical cancer?

- Television
- Radio
- Newspaper
- Health professional
- Relatives
- Friends
- FCHV
- Health related books and posters
- Internet based applications
3. Knowledge about cervical cancer: (source: https://www.ncbi.nlm.nih.gov/pmc/articles/PMC3996269/ https://bmcpublichealth.biomedcentral.com/articles/10.1186/s12889-018-5958-8) Yes No Don’t know
  a. Is cervical cancer Preventable? Can cervical cancer be a terminal illness?
  b. Is having multiple sexual partners a risk factor?
  c. Is smoking a risk factor for cervical cancer?
  d. Is oral contraceptive a risk factor for Cervical cancer?
  e. Is giving birth to too many babies a risk factor for Cervical cancer?
  f. Can cervical cancer be associated with an infection (HPV)?
  g. Are you more likely to get cervical cancer if your family has it?
  h. Is there an effective method that significantly reduces the risk of cervical cancer?
4. Do you think the given factor can reduce the risk of getting cervical cancer? (source: https://www.ncbi.nlm.nih.gov/pmc/articles/PMC3996269/) Yes No Don’t know
  a. A diet rich in so-called antioxidants?
  b. Regular physical exercise?
  c. Use of vitamin supplement?
  d. Proper long and relaxing sleep?
  e. Avoiding genetically modified food?
  f. Avoiding Highly processed food?
  g. Weight loss?
  h. Restraining from casual sex? **HPV Vaccine** (source: https://www.ncbi.nlm.nih.gov/pmc/articles/PMC3996269/) Yes No Don’t know

1. Have you ever heard about a vaccine against cervical cancer?
5. Where or from whom did you hear about the HPV vaccine?

- Television
- Radio
- Newspaper
- Health professional
- Relatives
- Friends
- FCHV
- Health related books and posters
- Internet based applications
2.
3. Is it available in Nepal?
4. Does it guarantee 100% protection from cervical cancer?
5. Do you know where you can get vaccinated?
6. Is it free of charge (covered by government insurance or immunization package)?
7. Have you ever been vaccinated?
8. What is the best age to get vaccinated?

- 8 years
- 9-13 years
- 14 - 18 years
- 19-25 years
- >25 years
- Don’t Know

**Cervical Cancer Screening (source:** https://www.who.int/ncds/surveillance/data-toolkit-for-cervical-cancer-prevention-control/en/)

6. Have you ever heard about Cervical cancer screening? Yes No don’t know
7. Where or from whom did you hear about cervical cancer screening?

- Television
- Radio
- Newspaper
- Health professional
- Relatives
- Friends
- FCHV
- Health related books and posters
- Internet based applications

“Now I’m going to ask you about tests a health-care worker can do to check for cervical cancer. The tests a health-care worker can do to check for cervical cancer are called a Pap smear, HPV test, and VIA test. Pap smear: “For a Pap smear test, a health-care worker puts a small stick or swab inside the vagina to wipe the cervix, and sends the sample to the laboratory.” HPV test: “For an HPV test, a small stick or swab is put inside the vagina to wipe the cervix, and the sample is sent to the laboratory. This can be done by a health-care provider or by a woman herself.” VIA: “For a VIA test, a health-care worker puts vinegar on the cervix and looks to see if the cervix changes color.”

1. Has a health-care worker ever tested you for cervical cancer? Yes No
2. At what age when you first tested for cervical cancer?
3. When did you have your last cervical cancer screening test?

- Less than 1 year ago
- 1–2 years ago
- 3–5 years ago
- More than 5 years ago
- Don’t know
- Refused
4. What cervical screening method was used in the last screen?

- HPV testing
- Pap Smear
- VIA test
- Others specify
- Don’t Know
- Refused
5. What is the MAIN reason you had your last test for cervical cancer?

- Part of routine examination
- Follow up on abnormal or inconclusive result
- Recommended by health-care provider
- Recommended by other source
- Experiencing pain or other symptoms
- Others
- Don’t know
- Refused

a. Others Specify
6. Where did you receive your last test for cervical cancer?

- Health post/Health Center
- Mobile camp/clinic
- Government hospital
- Private Hospital/ clinic
- Other Specify?
7. What was the result of your last test for cervical cancer?

- Did not receive result
- Normal/negative
- Abnormal/positive
- Suspect cancer Inconclusive
- Don’t know
- Refused
8. Did you have any follow-up visits because of your last test result? Yes No Don’t Know Refused
9. Did you have any treatment to your cervix because of your last test result? Yes No Don’t Know Refused
10. Did you receive the treatment for your cervix during the same visit as your last test for cervical cancer? Yes No Don’t Know Refused
11. What is the MAIN reason you did not receive treatment as a result of your last test result?

- Was not told I needed treatment
- Did not know how/where to get treatment
- Embarrassment
- Too expensive
- Didn’t have time
- Clinic too far away
- Poor service quality
- Afraid of the procedure
- Afraid of social stigma
- Cultural beliefs
- Family member would not allow it
- Other specify
- Don’t know
- Refused

a. Specify the relationship of the member to the respondent who did not allow
b. Specify other reason
12. What is the MAIN reason you have never had a cervical cancer test?

- Did not know how/where to get treatment
- Embarrassment
- Too expensive
- Didn’t have time
- Clinic too far away
- Poor service quality
- Afraid of the procedure
- Afraid of social stigma
- Cultural beliefs
- Family member would not allow it
- Other specify
- Don’t know
- Refused

a. Specify the relationship of the member to the respondent who did not allow
b. If others, specify
13. Would you be willing to collect a sample by yourself to test for cervical cancer in your home, if you were given instructions on how to collect the sample? Yes No Don’t Know Refused

## Notes

### Competing Interest Statement

The authors have declared no competing interest.

### Funding Statement

This study is funded by the National Cancer Institute.

### Author Declarations

The project has received IRB approval from the Ethical Review Board, Nepal Health Research Council (Jan 2020 /ref no 1699) and IRB at Icahn School of Medicine at Mount Sinai (Study-23-00362).

